# Seroconversion rate after primary vaccination with two doses of BNT162b2 versus mRNA-1273 in solid organ transplant recipients: a systematic review and meta-analysis

**DOI:** 10.1101/2021.12.23.21268314

**Authors:** Arno Verleye, Veerle Wijtvliet, Steven Abrams, Rachel Hellemans, Rania Bougrea, Annick Massart, Lissa Pipeleers, Karl Martin Wissing, Benedicte Y. De Winter, Pierre Van Damme, Daniel Abramowicz, Kristien J. Ledeganck

**Affiliations:** Department of Nephrology and Hypertension, Antwerp University Hospital, Edegem, Belgium; Global Health Institute, Family Medicine and Population Health, University of Antwerp, Wilrijk, Belgium; Data Science Institute, Interuniversity Institute for Biostatistics and statistical Bioinformatics, UHasselt, Diepenbeek, Belgium; Laboratory of Experimental Medicine and Pediatrics and member of the Infla-Med Centre of Excellence, University of Antwerp, Antwerp, Belgium; Centre for the Evaluation of Vaccination, Vaccine and Infectious Disease Institute, University of Antwerp, Antwerp, Belgium; Department of Nephrology, Universitair Ziekenhuis Brussel, Vrije Universiteit Brussel, Brussels, Belgium

## Abstract

In the general population, the seroconversion rate after primary vaccination with two doses of anti-SARS-CoV-2 mRNA vaccine reaches nearly 100%, with significantly higher antibody titers after mRNA-1273 vaccination compared to BNT162b2 vaccination. Here, we performed a systematic review and meta-analysis to compare the antibody response after two-dose mRNA-1273 versus BNT162b2 vaccination in solid organ transplant (SOT) recipients. A systematic literature research was performed in Pubmed, Web of Science, and the Cochrane library and original research papers were included for a meta-analysis to calculate vaccine-specific seroconversion rates for each of the mRNA vaccines. Next, the pooled relative seroconversion rate was estimated. Six studies that described the development of antibodies against receptor-binding domain (RBD) and/or S1 subunit of the spike protein were eligible for meta-analysis. Two of them also reported antibody titers. The meta-analysis revealed lower seroconversion rates in SOT recipients vaccinated with two doses of BNT162b2 (45.2%; 95% confidence interval (CI) 32.5%-58.3%) than patients vaccinated with two doses of mRNA-1273 (60.4%; 95% CI 47.4%-72.7%. The relative seroconversion rate amounted 0.79 (95% CI 0.71-0.88). This systematic review and meta-analysis indicates that, in SOT recipients, higher seroconversion rates were observed after vaccination with mRNA-1273 compared to BNT162b2.

## 1. Introduction

The COVID-19 pandemic rages since more than 1.5 years now. It is estimated that more than 200 million people have been infected, and that close to 5 million individuals have died (1). Solid organ transplant (SOT) patients carry a greater risk of complications or mortality attributable to SARS-CoV-2 infection (2). Therefore, an efficient vaccination strategy is critical in this population.

Both BNT162b2 and mRNA-1273 vaccines have each shown more than 90% efficacy in preventing COVID-19 illness in the general population (3, 4). As patients on immunosuppressive drugs were excluded from Phase III trials, little is known about the efficacy of these vaccines in SOT recipients.

Multiple reports show that in this latter group, only around 50% of patients develop anti-SARS-CoV-2 antibodies after a primary vaccination with two injections (5, 6).

Although both vaccines induce a nearly 100% seroconversion rate in the general population, several studies demonstrated significantly higher antibody titers after mRNA-1273 vaccination compared to BNT162b2 vaccination (7-10). In this systematic review and meta-analysis, we aimed to investigate 1) the proportion of SOT patients developing a humoral response to both vaccines as well as 2) the corresponding anti-SARS-CoV-2 spike antibody levels by performing a systematic review and meta-analysis of the existing literature. Higher seroconversion rates and/or antibody titers following either mRNA vaccine could potentially affect vaccination strategies targeting this vulnerable group.

## 2. Methods

A clinical research question was formulated according to the following PICO (Population, Intervention, Comparison, Outcome) (11): in SOT patients (P), do two doses of mRNA-1273 vaccination (I), compared to two doses of BNT162b2 vaccination (C), result in a higher seroconversion rate (O1) and/or higher anti-SARS-CoV-2 antibody titers (O2)?

Studies in which results are reported on both the antibody response after two doses of BNT162b2 (Pfizer/BioNTech, New York, United States) and mRNA-1273 (Moderna, Cambridge, Massachusetts, United States) in SOT recipients were considered eligible. Randomized controlled trials, cohort studies, case–control studies and cross-sectional studies were included. The search period was limited from 2020 to 2021. No age restriction was applied. Literature reviews, case reports and commentaries were excluded.

A systematic search of three databases was conducted (PubMed, Web of Science and Cochrane library) using the following search terms: transplant* AND vaccin* AND (mRNA OR Moderna OR Pfizer BioNTech OR mRNA-1273 OR BNT162b2 OR Comirnaty OR Spikevax). The last search date was 26/09/2021. In addition, we included data from our own kidney transplant population, as recently published as a preprint to medRxiv (12).

To minimize selection bias, studies were screened independently by two reviewers (A.V. and R.B.). First, duplicates were removed, after which articles were screened by title and abstract. Remaining reports were subsequently assessed for eligibility through full text screening. Finally, the methodological quality of the included studies was assessed using the Methodological Index for Non-Randomized Studies (MINORS) (13). Indeed, we could not retrieve any randomized controlled trial on this topic. Any disagreement was resolved by consensus.

A.V. extracted the following data from the included studies: cohort size, transplant type, seroconversion rate, antibody titer, immunological assay and time of measurement. A second author (K.J.L.) checked the data for correctness.

The meta-analysis was performed using the packages *metafor and meta* in the statistical software package R, version 4.1.2 (14). More specifically, a single group random-effects meta-analysis approach was considered to pool the seroconversion rates for each of the mRNA vaccines (mRNA-1273 and BNT162b2) obtained from the eligible studies. A Freeman-Tukey double arcsine transformation of the study-specific seroconversion rates and corresponding standard errors were used in the pooling procedure. The inverse-variance method was used to weight the study-specific transformed seroconversion rates (with the inverse of the within-study variance as study-specific weights). The between-study variability was estimated using the DerSimonian-Laird estimator. Heterogeneity across the studies was quantified by means of the inconsistency index or I^2^-statistic (Higgins and Thompson, 2002; Higgins *et al*., 2003). Next to the single-group meta-analysis models for each of the mRNA vaccines, we performed a random-effects meta-analysis of the relative seroconversion rates for the two two-dose mRNA vaccination schemes in SOT patients. Again, the inverse-variance method and DerSimonian-Laird estimator were used.

## 3. Results

Our search yielded a total of 355 results (Pubmed N = 223, Web of Science N = 128 and Cochrane library N = 4). After removal of duplicates and screening by title/abstract, 34 articles were found to be eligible for full-text reading. Of these, 29 articles were excluded, mostly because no comparison was made between both mRNA vaccines. After adding the results of our own research (12), a total of 6 studies was included in this meta-analysis. The full study selection process is shown in Figure 1.

**Figure 1.**
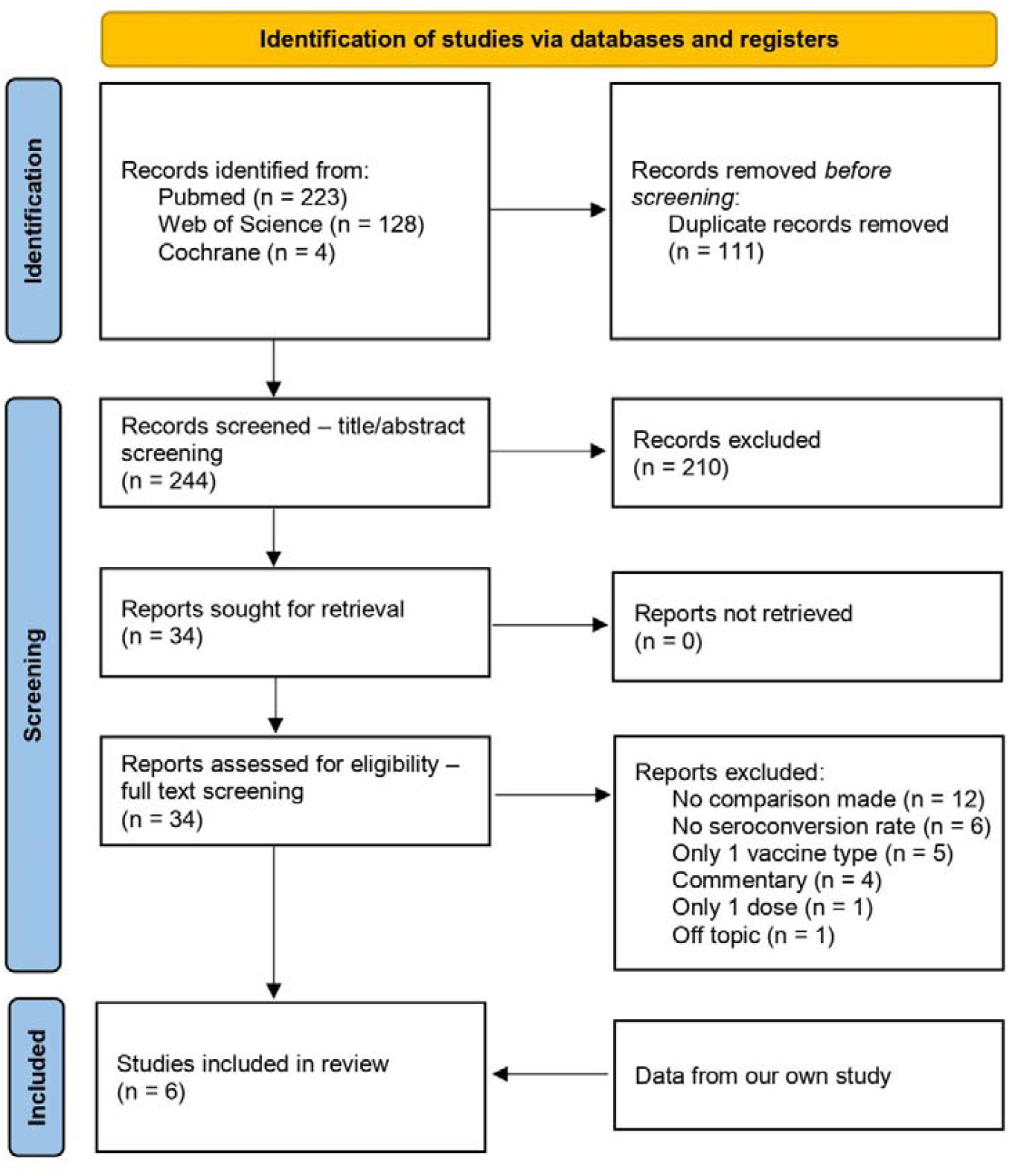
Flow diagram of study selection From: Page MJ, McKenzie JE, Bossuyt PM, Boutron I, Hoffmann TC, Mulrow CD, et al. The PRISMA 2020 statement: an updated guideline for reporting systematic reviews. BMJ 2021;372:n71. doi: 10.1136/bmj.n71

### 3.1 Study characteristics

Study characteristics are summarized in Table 1. All studies were prospectively designed. There was a considerable heterogeneity in transplant type as well as in the immunological assay that was used to measure antibodies. Among the 1630 SOT patients studied, 823 were transplanted with a kidney, 290 with a liver, 247 with a lung, 231 with a heart, 5 with a pancreas, and 20 SOT were multi-organ recipients. The vaccine specific seroconversion rate was available from 1586 patients. A total of 791 patients was vaccinated with BNT162b, 795 patients received the mRNA-1273 vaccine. The anti-SARS-Cov 2 antibodies against receptor binding domain (RBD) and/or S1 subunit of the spike protein were detected with the Roche-Elecsys (anti-RBD) or the Euroimmun test (anti-S1) in 3 studies (15-17), with only the Euroimmun test in 1 study (18), with the Abbot Alimiti (anti-RBD) in 1 study (19), and with a locally-designed Luminex platform (anti-RBD) in 1 study (12). The median time of the antibody response varied between 17 days and 5 weeks after second vaccine administration.

**Table 1.** Study characteristics

| Author (year) | Study design | SOT recipients | Transplant type (N) | BNT162b2 vaccinated | BNT162b2 seroconverted | mRNA-1273 vaccinated | mRNA-1273 seroconverted | Immunological assay | Cutoff seroconversion | Median time of measurement |
| --- | --- | --- | --- | --- | --- | --- | --- | --- | --- | --- |
| Boyarsky (2021) | Prospective cohort study | 658* | Kidney (322)<br>Liver (129)<br>Heart (97)<br>Lung (71)<br>Pancreas (5)<br>Multiorgan (26) | 342 | 167 | 307 | 183 | Roche Elecsys (anti-RBD total Ab) OR EUROIMMUN (anti-S1 IgG) | ≥ 0,8 U/ml (Roche Elecsys)<br>≥ 1:1 arbitrary units (EUROIMMUN) | 29 days |
| Hallett (2021) | Prospective cohort study | 237 | Heart (134) | 70 | 42 | 64 | 41 | Roche Elecsys (anti-RBD total Ab) OR EUROIMMUN (anti-S1 IgG) | ≥ 0,8 U/ml (Roche Elecsys)<br>≥ 1:1 arbitrary units (EUROIMMUN) | 29 days |
|  |  |  | Lung (103) | 56 | 19 | 47 | 18 |  |  |  |
| Narasimhan (2021) | Prospective cohort study | 73 | Lung (73) | 48 | 9 | 25 | 9 | Abbott Alinity i (anti-RBD IgG) | ≥ 50 AU/mL | 17.5 days (BNT162b)<br>19 days (mRNA-1273) |
| Strauss (2021) | Prospective cohort study | 161 | Liver (161) | 85 | 62 | 76 | 68 | Roche Elecsys (anti-RBD total Ab) OR EUROIMMUN (anti-S1 IgG) | ≥ 0,8 U/ml (Roche Elecsys)<br>≥ 1:1 arbitrary units (EUROIMMUN) | 30 days |
| Stumpf (2021) | Prospective cohort study | 368* | Kidney (368) | 99 | 26 | 234 | 114 | EUROIMMUN (anti-S1 IgG OR anti-S1 IgA) | A positive serologic response was defined as de novo antibody development (seroconversion) | 4 to 5 weeks |
| Wijtvliet (2021) | Prospective cohort study | 133 | Kidney (133) | 91 | 51 | 42 | 32 | Luminex (anti-RBD IgG) | signal-to-noise ratio > 1 | 28 days |

\* vaccine specific seroconversion rate not available from all included patients

### 3.2 Risk of bias within studies

The MINORS criteria revealed a mean score of 75%. Three out of 6 studies were considered high quality (12, 16, 18), the other half was scored as moderate quality (15, 17, 19), as shown in Table 2. The risk of bias was thus acceptable.

**Table 2.** Individual MINORS score

|  | Boyarsky<br>2021 (15) | Hallett<br>2021 (16) | Narasimhan<br>2021 (19) | Strauss<br>2021 (17) | Stumpf<br>2021 (18) | Wijtvliet<br>2021 (12) |
| --- | --- | --- | --- | --- | --- | --- |
| Clearly stated aim | 2 | 2 | 2 | 2 | 2 | 2 |
| Inclusion of consecutive patients | 1 | 2 | 1 | 2 | 2 | 1 |
| Prospective data collection | 1 | 2 | 1 | 2 | 2 | 2 |
| Endpoints appropriate to study aim | 2 | 2 | 2 | 2 | 2 | 2 |
| Unbiased assessment of study endpoint | 0 | 0 | 0 | 0 | 0 | 2 |
| Follow-up period appropriate to study aim | 2 | 2 | 2 | 2 | 2 | 2 |
| < 5% loss to follow-up | 2 | 2 | 2 | 0 | 2 | 2 |
| Prospective calculation of study size | 2 | 2 | 2 | 2 | 2 | 2 |
| Adequate control group | 0 | 0 | 0 | 0 | 0 | 0 |
| Contemporary groups | 2 | 2 | 2 | 2 | 2 | 2 |
| Baseline equivalence of groups | 0 | 1 | 1 | 1 | 1 | 1 |
| Adequate statistical analyses | 2 | 2 | 2 | 2 | 2 | 2 |
| Total score | 16/24<br>(67%) | 19/24<br>(79%) | 17/24<br>(71%) | 17/24<br>(71%) | 19/24<br>(79%) | 20/24<br>(83%) |

The items are scored 0 (not reported), 1 (reported but inadequate), or 2 (reported and adequate).
The global ideal score is 24 for comparative studies. The corresponding scores are 0–6, very low quality; 7–12, low quality; 13–18, moderate quality; and 19–24, high quality.

### 3.3 Synthesis of results

No patients had prior polymerase chain reaction-confirmed diagnosis of COVID-19. Two studies screened for anti-nucleocapsid protein IgG prior to vaccination. While excluded by Stumpf *et al*., Narasimhan *et al*. included one patient with a past SARS-CoV-2 infection (18, 19).

The single group meta-analysis models indicated considerable heterogeneity across different studies, with high I^2^-values (91.2%, 95% CI: [84.5%, 95.0%] and 91.6%, 95% CI: [85.3%, 95.2%] for mRNA-1273 and BNT162b2, respectively). The pooled seroconversion rate was estimated to be higher for mRNA-1273 (60.4%, 95% CI: [47.4%, 72.7%]; Figure 2b) as compared to BNT162b2 (45.2%, 95% CI: [32.5%, 58.3%]; Figure 2a). As presented in Figure 3, the relative seroconversion rate was estimated to be 0.788 (95% CI: [0.707, 0.879]) for BNT162b2 vs. mRNA-1273 vaccination based on a random-effects meta-analysis model as described above. Consequently, a significantly lower seroconversion rate was observed after two-dose vaccination with BNT162b2 as compared to mRNA-1273.

**Figure 2.**
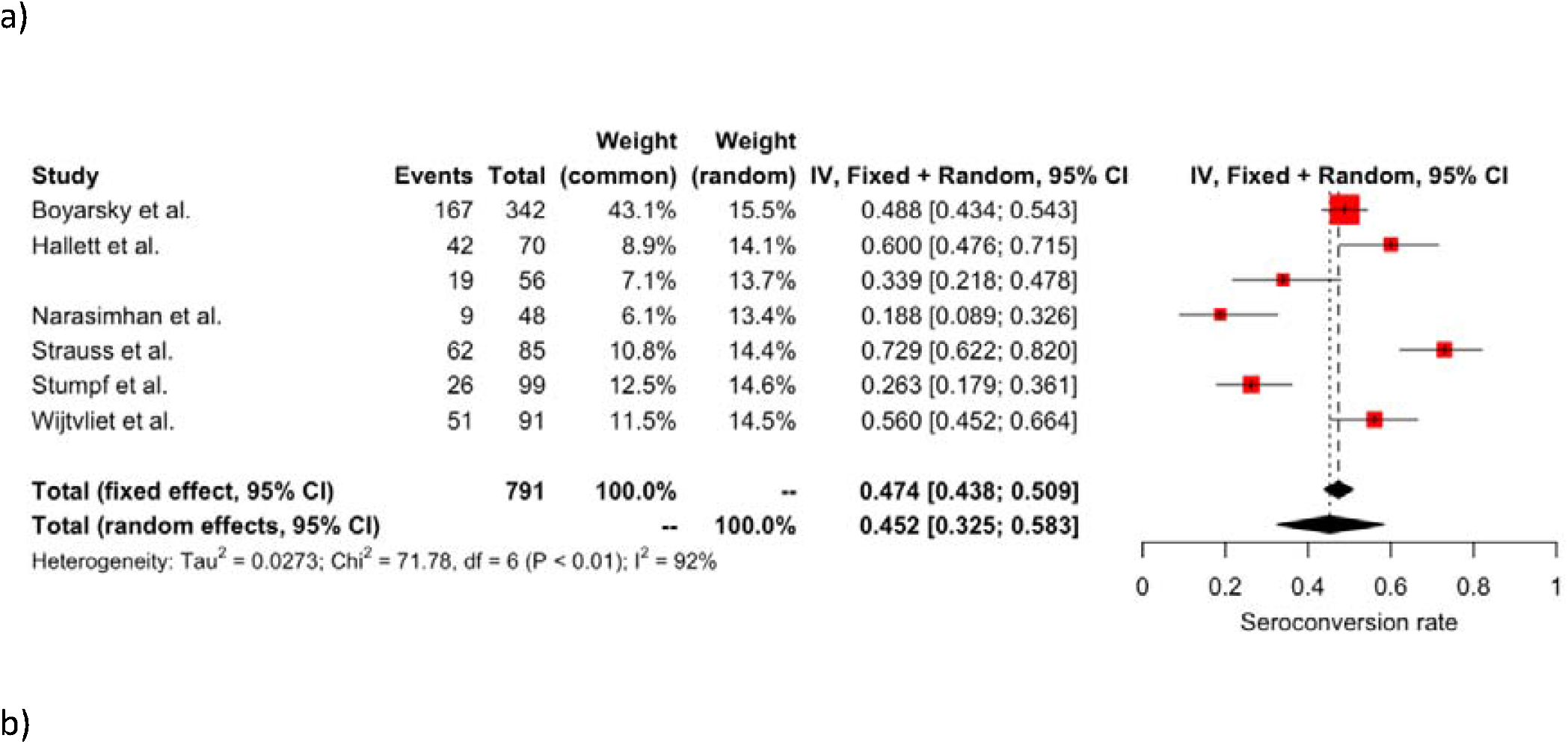
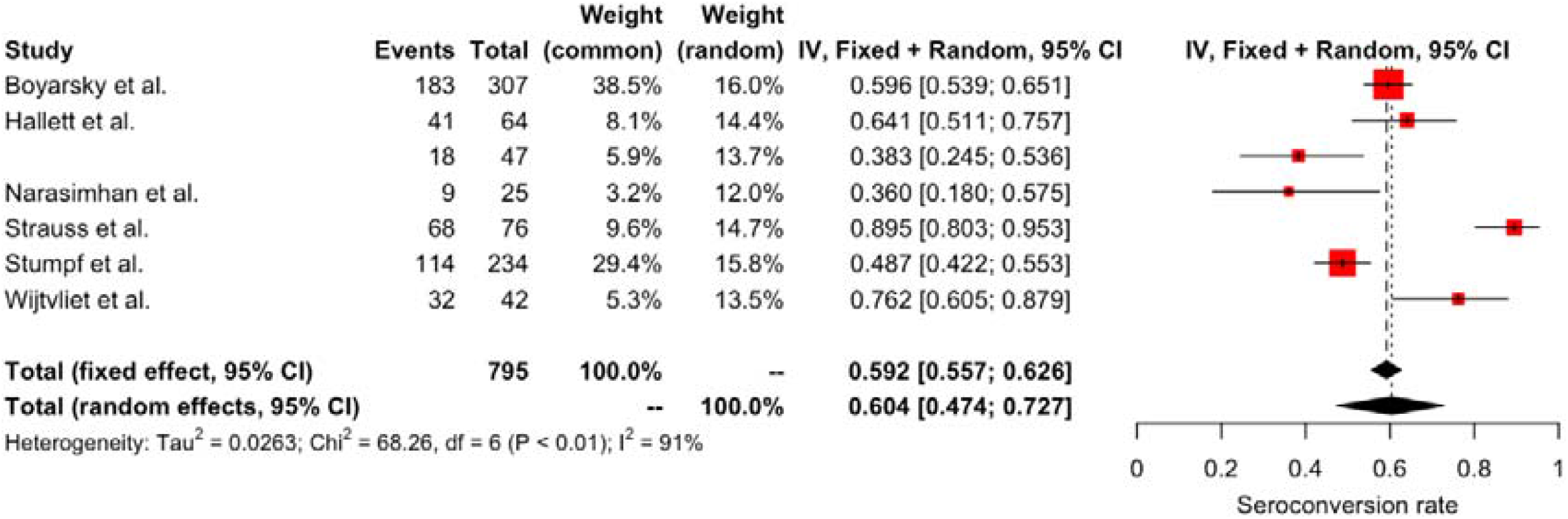
Study-specific and pooled estimates for the seroconversion rate after two-dose mRNA vaccination with BNT162b2 (panel a) or mRNA-1273 (panel b) based on random-effects meta-analysis models and relying on the inverse-variance (IV) method. Box sizes in the forest plots are proportional to the weight assigned to each study. Limits of the displayed intervals are defined as 95% confidence intervals (CIs). Six studies calculated the seroconversion rates in SOT recipients after two-dose BNT162b2 vaccination (n = 791), resulting in a pooled [95% CI] seroconversion rate of 45.2% [32.5%, 58.3%] (panel a). The same six studies also described seroconversion rates in SOT recipients after two-dose mRNA-1273 vaccination (n = 795), resulting in a pooled [95% CI] seroconversion rate of 60.4% [47.4%, 72.7%] (panel b). 95% CI = 95% Confidence Interval; df = degrees of freedom; I^2^ = inconsistency index; IV = inverse variance method.

**Figure 3.**
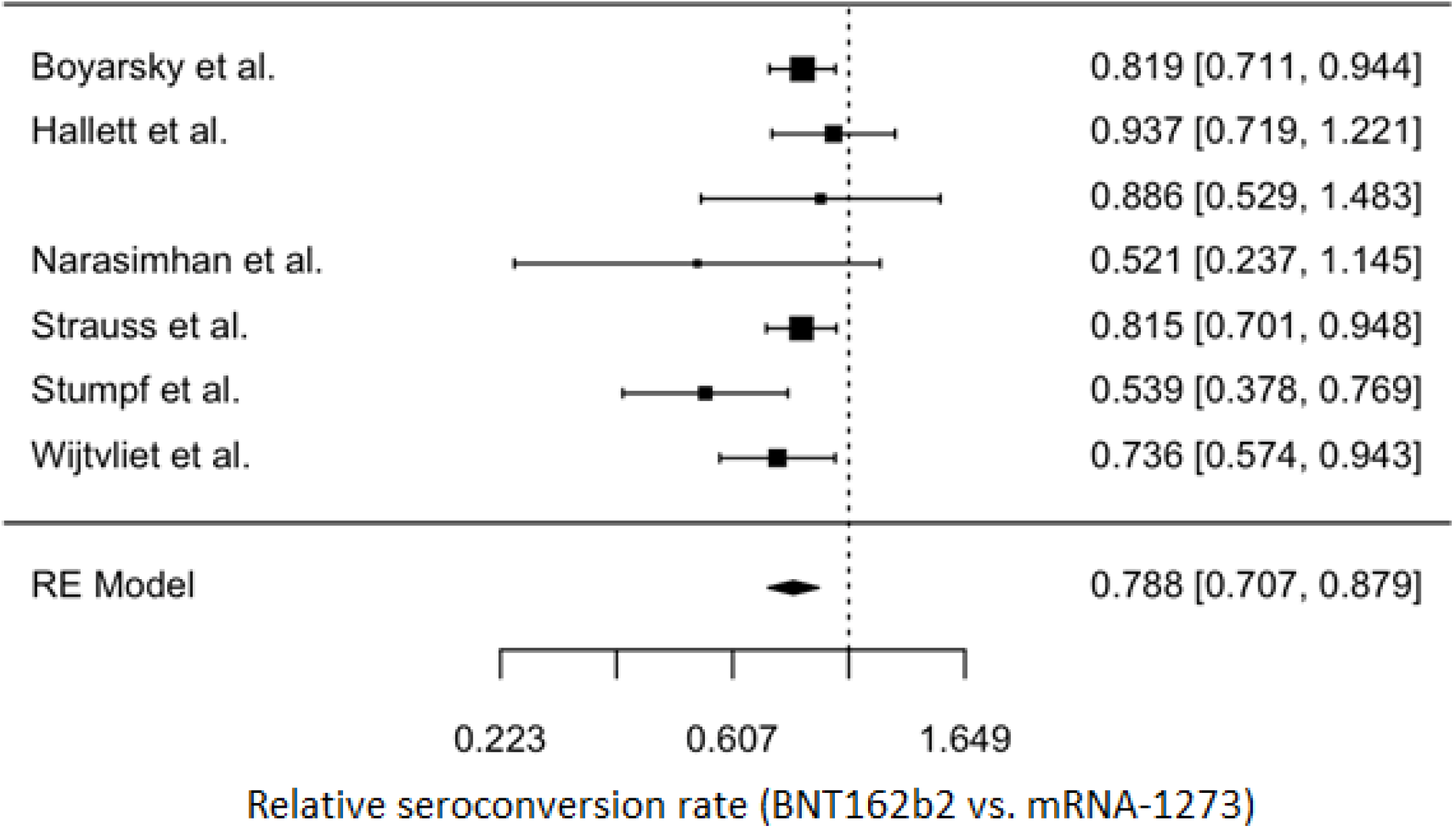
Meta-analytic result for the relative seroconversion rate (BNT162b2 vs. mRNA-1273) based on a random-effects meta-analysis model and relying on the inverse-variance method. Seroconversion rates appeared to be significantly lower in patients vaccinated with two doses of BNT162b2 than patients vaccinated with two doses of mRNA-1273 (78.8% [70.7%, 87.9%]). RE = random-effects.

Only two studies directly compared anti-SARS-CoV-2 antibody titers after vaccination with either mRNA vaccine (12, 19), therefore precluding to perform a meta-analysis on this issue. While Narasimhan et al. (19) did not find a significant difference in antibody titers between the BNT162b2 vaccine (median 0.9 AU/ml (95% CI 0.0-4.1)) and the mRNA-1273 formulation (median 20.6 AU/ml (95% CI 0.8-80.2)) among lung transplant patients (p = 0.96), Wijtvliet et al. (12) showed significantly higher antibody titers after two doses of mRNA-1273 compared to BNT162b2 in kidney transplant recipients (mean log-transformed antibody levels were 0.289 units higher for Moderna vs. Pfizer vaccination in multiple linear mixed model (p = 0.005)).

Two out of 6 studies reported on T-cell anti-SARS-CoV-2 response (18, 19). Interestingly, Stumpf et al. showed a numerically higher cellular immune response after vaccination with mRNA-1273 as compared to BNT162b2. Narasimhan *et al*. did not compare T-cell response between both mRNA vaccines, but studied the humoral response in relation to T-cell activity.

## 4. Discussion

This systematic review and meta-analysis reveal that, in SOT patients, vaccination with mRNA-1273 leads to a significantly higher seroconversion rate than BNT162b2 vaccination (60.4% vs. 45.2% respectively with a relative seroconversion rate of 0.79 (95% CI 0.71-0.88)). In a similar research in patients with hematologic malignancies, the seroconversion rate was 56% with mRNA-1273, versus 33% with BNT162b2 (p = 0.013) (20). This contrasts with dialysis patients, where the seroconversion rate is much higher for both the BNT162b2 (around 87-88%) (12, 21) and the mRNA-1273 vaccine (around 94.4-100%) (12, 21). In the paper by Lacson *et al*., no difference in seroconversion rate was observed between both vaccines in dialysis patients (p = 0.42) while in the study by Wijtvliet *et al*., mRNA-1273 led to a higher seroconversion rate (p = 0.005).

Among the studies included in this systematic review and meta-analysis, only two articles directly compared anti-SARS-CoV-2 antibody titers after vaccination with either mRNA vaccine (12, 19), therefore precluding to perform a meta-analysis on this issue. While Narasimhan *et al*. (19) did not find a significant difference in antibody titers between the BNT162b2 vaccine and the mRNA-1273 formulation among lung transplant patients (p = 0.95), Wijtvliet et al. (12) showed significantly higher antibody titers after two doses of mRNA-1273 compared to BNT162b2 in kidney transplant recipients. In the general population, whereas the seroconversion rates are similar between the two vaccines, there is now clear evidence that higher titers of anti-SARS-CoV-2 antibodies are present after vaccination with mRNA-1273 as compared to BNT162b2 (7-10). This can lead to serious clinical consequences. Indeed, although the incidence of severe or critical COVID-19 illness remains low in the fully vaccinated general population, a higher number of patients with severe or critical illness was observed in those who received the BNT162b2 vaccine than in those who received mRNA-1273 (22). In a case-control study of patients hospitalized for Covid-19, estimated vaccine effectiveness of both mRNA vaccines was similar within 120 days of vaccination. In contrast, beyond 120 days, a higher estimated effectiveness to prevent COVID19 hospitalizations was observed after vaccination with mRNA-1273 vs. BNT162b2 (85% vs. 64%, p < 0.01) (23).

The difference in immunogenicity between those two mRNA vaccines could relate to the amount of mRNA used in the respective vaccines. Indeed, the mRNA-1273 vaccine contains 100 μg of mRNA while the BNT162b2 vaccine only contains 30 μg. Another possible explanation is the longer interval between priming and boosting for mRNA-1273 (4 weeks as compared to 3 weeks for BNT162b2).

Furthermore, there are differences in the lipid composition of the nanoparticles used for packaging the mRNA content of mRNA-1273 and BNT162b2. BNT162b2 has a lipid nanoparticle composed of ALC-0315, ALC-0159, DSPC and cholesterol whereas the lipid nanoparticle of mRNA-1273 is composed of SM-102, PEG-DMG, DSPC and cholesterol (24).

This meta-analysis makes clear that the current research on immunity after SARS-CoV-2 vaccination in vulnerable patients has several limitations. First, given the fact that the response after vaccination against SARS-CoV-2 has only been investigated since less than a year, only 6 studies, reporting on 1586 patients could be included into this systematic review and meta-analysis. However, even with this restrictive number of papers, the results were consistent across all studies making this meta-analysis sound. Second, the included studies were all observational in nature; none of them was a randomized controlled trial. Third, the number of studies reporting on vaccine specific antibody titers was too small to allow for a meta-analysis, and only two studies reported on T-cell response. Finally, there was a considerable amount of heterogeneity across the different studies, which could be explained by the different transplant types analyzed. However, this heterogeneity disappeared when looking at the relative differences across patient groups (cfr. I^2 value: 2% of total variability due to between-study variability). Larger studies with stratification by age, gender, transplant type and immunosuppressive drugs could help overcome this problem.

In conclusion, the seroconversion rate appeared to be higher after mRNA-1273 vaccination vs. BNT162b2 vaccination in SOT recipients. Future studies are needed to assess whether these differences are also associated with a better clinical protection against severe disease, hospitalization and/or mortality. This will help to decide whether mRNA-1273 should be the preferred vaccine in SOT recipients. In addition, all efforts should be made to vaccinate kidney transplant candidates before transplantation, as the overall efficacy of SARS-CoV-2 vaccines is better during dialysis than after kidney transplantation (25).

## Data Availability

All data produced in the present study are available upon reasonable request to the authors

## Abbreviations

CI: confidence interval
df: degrees of freedom
I2: inconsistency index
IV: inverse variance
method
RBD: receptor-binding domain
RE: random-effects
SOT: solid organ transplant

## Author Contributions

AV and RB performed the literature search independently, assessed the methodological quality, and AV subsequently extracted data from the articles that met the inclusion criteria. KJL checked the data for correctness. SA performed the meta-analysis. AV, RH, VW, KJL, and DA drafted the manuscript. AV, VW, SA, RH, AM, LP, KMW, BDW, PVD, DA and KJL reviewed and approved the final version of the manuscript.

## Acknowledgments

The authors would like to thank Erik Snelders for his outstanding secretarial help.

## Data availability statement

The data that support the findings of this study are available from the corresponding author upon reasonable request.

## References

1. Organization WH. World Health Organization coronavirus disease (COVID-19) dashboard. World Health Organization 2020.

2. Azzi Y, Bartash R, Scalea J, Loarte-Campos P, Akalin E. COVID-19 and Solid Organ Transplantation: A Review Article. Transplantation 2021;105(1):37–55.

3. Polack FP, Thomas SJ, Kitchin N, Absalon J, Gurtman A, Lockhart S et al. Safety and Efficacy of the BNT162b2 mRNA Covid-19 Vaccine. New England Journal of Medicine 2020;383(27):2603–2615.

4. Baden LR, El Sahly HM, Essink B, Kotloff K, Frey S, Novak R et al. Efficacy and Safety of the mRNA-1273 SARS-CoV-2 Vaccine. New England Journal of Medicine 2020;384(5):403–416.

5. Marinaki S, Adamopoulos S, Degiannis D, Roussos S, Pavlopoulou ID, Hatzakis A et al. Immunogenicity of SARS-CoV-2 BNT162b2 vaccine in solid organ transplant recipients. Am J Transplant 2021;21(8):2913–2915.

6. Marion O, Del Bello A, Abravanel F, Couat C, Faguer S, Esposito L et al. Safety and Immunogenicity of Anti-SARS-CoV-2 Messenger RNA Vaccines in Recipients of Solid Organ Transplants. Ann Intern Med 2021.

7. Steensels D, Pierlet N, Penders J, Mesotten D, Heylen L. Comparison of SARS-CoV-2 Antibody Response Following Vaccination With BNT162b2 and mRNA-1273. JAMA 2021.

8. Markewitz R, Pauli D, Dargvainiene J, Steinhagen K, Engel S, Herbst V et al. The temporal course of T-and B-cell-responses to vaccination with BNT162b2 and mRNA-1273. Clin Microbiol Infect 2021:S1198-1743X(1121)00496-00491.

9. Richards NE, Keshavarz B, Workman LJ, Nelson MR, Platts-Mills TAE, Wilson JM. Comparison of SARS-CoV-2 Antibody Response by Age Among Recipients of the BNT162b2 vs the mRNA-1273 Vaccine. JAMA Netw Open 2021;4(9):e2124331–e2124331.

10. Self WH, Tenforde MW, Rhoads JP, Gaglani M, Ginde AA, Douin DJ et al. Comparative Effectiveness of Moderna, Pfizer-BioNTech, and Janssen (Johnson & Johnson) Vaccines in Preventing COVID-19 Hospitalizations Among Adults Without Immunocompromising Conditions - United States, March-August 2021. MMWR Morb Mortal Wkly Rep 2021;70(38):1337–1343.

11. Richardson WS, Wilson MC, Nishikawa J, Hayward RS. The well-built clinical question: a key to evidence-based decisions. ACP J Club 1995;123(3):A12–13.

12. Wijtvliet VPWM, Ariën KK, Abrams S, Couttenye MM, Mestrez F, Mariën J et al. mRNA-1273 vaccine (Moderna): a better option than BNT162b2 (Pfizer) in kidney transplant recipients and dialysis patients? medRxiv 2021:2021.2009.2015.21263320.

13. Slim K, Nini E, Forestier D, Kwiatkowski F, Panis Y, Chipponi J. Methodological index for non-randomized studies (minors): development and validation of a new instrument. ANZ J Surg 2003;73(9):712–716.

14. Team RC. R: A Language and Environment for Statistical Computing. R Foundation for Statistical Computing 2018.

15. Boyarsky BJ, Werbel WA, Avery RK, Tobian AAR, Massie AB, Segev DL et al. Antibody Response to 2-Dose SARS-CoV-2 mRNA Vaccine Series in Solid Organ Transplant Recipients. JAMA 2021;325(21):2204–2206.

16. Hallett AM, Greenberg RS, Boyarsky BJ, Shah PD, Ou MT, Teles AT et al. SARS-CoV-2 messenger RNA vaccine antibody response and reactogenicity in heart and lung transplant recipients. J Heart Lung Transplant 2021.

17. Strauss AT, Hallett AM, Boyarsky BJ, Ou MT, Werbel WA, Avery RK et al. Antibody response to SARS-CoV-2 messenger RNA vaccines in liver transplant recipients. Liver Transpl 2021.

18. Stumpf J, Siepmann T, Lindner T, Karger C, Schwöbel J, Anders L et al. Humoral and cellular immunity to SARS-CoV-2 vaccination in renal transplant versus dialysis patients: A prospective, multicenter observational study using mRNA-1273 or BNT162b2 mRNA vaccine. Lancet Reg Health Eur 2021:100178.

19. Narasimhan M, Mahimainathan L, Clark AE, Usmani A, Cao J, Araj E et al. Serological Response in Lung Transplant Recipients after Two Doses of SARS-CoV-2 mRNA Vaccines. Vaccines (Basel) 2021;9(7).

20. Ollila TA, Lu S, Masel R, Zayac A, Paiva K, Rogers RD et al. Antibody Response to COVID-19 Vaccination in Adults With Hematologic Malignant Disease. JAMA Oncol 2021;7(11):1714–1716.

21. Lacson E, Argyropoulos CP, Manley HJ, Aweh G, Chin AI, Salman LH et al. Immunogenicity of SARS-CoV-2 Vaccine in Dialysis. Journal of the American Society of Nephrology 2021;32(11):2735–2742.

22. Juthani PV, Gupta A, Borges KA, Price CC, Lee AI, Won CH et al. Hospitalisation among vaccine breakthrough COVID-19 infections. Lancet Infect Dis 2021;21(11):1485–1486.

23. Tenforde MW, Self WH, Adams K, Gaglani M, Ginde AA, McNeal T et al. Association Between mRNA Vaccination and COVID-19 Hospitalization and Disease Severity. JAMA 2021.

24. Schoenmaker L, Witzigmann D, Kulkarni JA, Verbeke R, Kersten G, Jiskoot W et al. mRNA-lipid nanoparticle COVID-19 vaccines: Structure and stability. Int J Pharm 2021;601:120586.

25. Quiroga B, Soler MJ, Ortiz A, Vaquera SM, Mantecón CJJ, Useche G et al. Safety and immediate humoral response of COVID-19 vaccines in chronic kidney disease patients: the SENCOVAC study. Nephrol Dial Transplant 2021.

